# Erythrocyte Osmotic Fragility as A Diagnostic Marker in Glaucoma: A Comprehensive Analysis Using LASSO Regression

**DOI:** 10.1101/2024.09.05.24312988

**Authors:** Jialiang Yang, Fang Yang, Junming Gu, Yilian Cheng, Qian Luo, Fang Hao, Bo Gong, Houbin Zhang

## Abstract

**Objective:** This study investigates the potential of blood biomarkers in the early diagnosis of glaucoma, focusing on erythrocyte osmotic fragility (EOF) as a novel indicator. We used Least Absolute Shrinkage and Selection Operator (LASSO) regression to evaluate EOF’s predictive utility for glaucoma diagnosis.

**Methods:** We included 195 confirmed glaucoma patients and an equal number of age- and sex-matched healthy controls. Blood samples were analyzed for various parameters, including white blood cell count, neutrophil count, red blood cell (RBC) count, and EOF. Key differential markers were identified, and a predictive model was constructed using LASSO regression models.

**Results:** LASSO regression analysis identified HCT, NEUT, LYMPH, MCV, MCHC, and EOF as critical blood biomarkers discriminating glaucoma patients from healthy controls. Incorporating EOF into the model significantly enhanced its predictive performance, with EOF showing a positive correlation with the likelihood of glaucoma.

**Conclusions:** EOF is a promising predictive biomarker for glaucoma. Combining EOF with other blood biomarkers significantly improves the accuracy of glaucoma diagnosis.

## Introduction

Glaucoma is the leading cause of irreversible vision loss worldwide, characterized by complex and multifactorial pathophysiology involving both known and potential risk factors [1–4]. Although glaucoma is traditionally linked to elevated intraocular pressure (IOP), clinical evidence suggests that approximately 30% of glaucoma patients exhibiting normal IOP (<21 mmHg) still suffer from irreversible optic nerve damage [5]. This observation implies the presence of additional critical pathogenic factors that the underlie pathophysiology of glaucoma beyond IOP [6].

Recent advances in molecular biology have highlighted the potential role of blood biomarkers in the onset and progression of glaucoma [7–11]. Blood biomarkers not only provide a systemic reflection of physiological and pathological status but also reveal specific pathological alterations in glaucoma patients, such as oxidative stress and inflammation [8, 12–15]. These alterations may play a significant role in the pathophysiology of glaucoma.

Oxidative stress and inflammation are central elements in the pathogenesis of glaucoma [10, 12, 15–17]. Oxidative stress elevates reactive oxygen species (ROS) levels, directly damaging erythrocyte membranes by inducing lipid peroxidation and reducing membrane stability [15, 18–22]. Inflammation exacerbates this damage by releasing inflammatory mediators (e.g., TNF-α, IL-1β), further compromising erythrocyte membrane integrity. As erythrocytes are primarily responsible for oxygen transport, increased erythrocyte osmotic fragility (EOF) affects oxygen transport mainly by causing cell membrane rupture, shape changes, and reduced elasticity [23–28]. This makes red blood cells (RBCs) more prone to rupture in the microcirculation, decreasing the number of functional RBCs. Additionally, membrane fragility impacts membrane protein function and disrupts hemoglobin’s ability to bind and release oxygen, thereby reducing overall oxygen transport efficiency. [23–28]. Therefore, EOF may reflect membrane stability and indicate ongoing pathological processes in glaucoma, such as chronic oxidative stress and inflammation.

Therefore, we hypothesize that elevated EOF is associated with glaucoma. Based on this hypothesis, we propose that EOF, as a novel biomarker, can effectively differentiate glaucoma patients from healthy controls, thereby improving the accuracy of early glaucoma diagnosis

This study aims to systematically evaluate the diagnostic potential of EOF and other blood biomarkers in glaucoma using the Least Absolute Shrinkage and Selection Operator (LASSO) [29–32] regression model. By constructing an efficient diagnostic model, we seek to provide a robust theoretical foundation and practical guidance for early glaucoma screening and precise diagnosis.

## Materials and Methods

### 1. Study Population

This study included 195 confirmed glaucoma patients and 195 age- and sex-matched healthy controls. All participants were provided informed consent, and the study was conducted following the Declaration of Helsinki and approved by the ethics committees of Sichuan Provincial People’s Hospital.

Participants in the glaucoma group met the International Glaucoma Association’s diagnostic criteria, including IOP ≥21 mmHg, structural optic nerve changes (e.g., optic atrophy, increased cup-to-disc ratio), and visual field defects. Patients with other severe ocular diseases, recent ocular trauma, or significant systemic conditions were excluded.

The control group was selected based on comprehensive ophthalmological examinations confirming the absence of glaucoma or other major ocular diseases, with age and sex matched to the glaucoma group. Exclusion criteria included systemic diseases affecting ocular health, recent ocular surgery, and medications influencing IOP.

### 2. Blood Sample Collection and Analysis

Fasting peripheral blood samples were collected in the morning using EDTA tubes (BD, USA) by trained technicians. Samples were stored at 4°C and analyzed within two hours. Blood samples were analyzed using an automated hematology analyzer (Mindray, China) to measure parameters such as white blood cell count (WBC), neutrophils (NEUT), lymphocytes (LYMPH), monocytes (MONO), eosinophils (EOS), basophils (BASO), RBC count, hemoglobin (HGB), hematocrit (HCT), mean corpuscular volume (MCV), mean corpuscular hemoglobin (MCH), mean corpuscular hemoglobin concentration (MCHC), red cell distribution width-standard deviation (RDW-SD), and platelet count (PLT).

### 3. EOF test (EOFT)

EOF was measured using a standardized osmotic fragility test [33], which assesses erythrocyte membrane stability by determining the critical osmotic pressure at which erythrocytes rupture. Briefly, 50 µL of blood was added to different concentrations of hypotonic saline solutions containing0.7%, 0.65%, 0.6%, 0.55%, 0.5%, 0.45%, 0.4%, 0.35%, 0.3%, or 0.25% NaCl (Sigma, USA) and gently mixed. The solutions were left at room temperature (25°C) for 2 hours. Afterward, the red blood cells’ resistance to the hypotonic solutions was examined by assessing hemolysis. The concentration of the hypotonic saline solution at which hemolysis first began indicated the minimum resistance of the red blood cells in the blood, while complete hemolysis indicated the maximum resistance. Lower resistance to hypotonic saline solutions signifies greater fragility of the red blood cells, whereas higher resistance indicates less fragility. The range from maximum resistance to minimum resistance is referred to as the fragility range.

### 4. Giemsa staining

Giemsa staining is a classic staining method used to observe the morphology of blood cells [34]. Blood was used to prepare a thin smear and allowed for air dry. The smear was fixed with methanol for 5 minutes. The A Giemsa staining kit (Yeasen, China) was used to stain the smear follow the manufacturer’s instruction. The stained smear was imaged under a microscope.

### 5. Statistical Analysis

LASSO regression (Least Absolute Shrinkage and Selection Operator) is a linear regression method that introduces an L1 regularization term to handle high-dimensional data and multicollinearity issues. It can shrink the coefficients of unimportant features to zero, thereby achieving feature selection and model sparsity. LASSO regression not only improves the predictive performance of models but also enhances their interpretability, making it an effective tool for analyzing high-dimensional datasets, with widespread applications across various fields.

LASSO regression introduces an L1 regularization term in the regression model to handle multicollinearity effectively and automatically select the most useful features for prediction [29–32]. All features were standardized, and cross-validation was used to select the optimal regularization parameter λ. After fitting the LASSO model on the training set, features with non-zero coefficients were extracted to construct the final predictive model. Model performance was evaluated through independent validation sets. Sensitivity analyses were conducted to verify the model’s applicability and robustness across different subgroups (e.g., age, sex) (**Figure 1A**).

**Figure 1.**
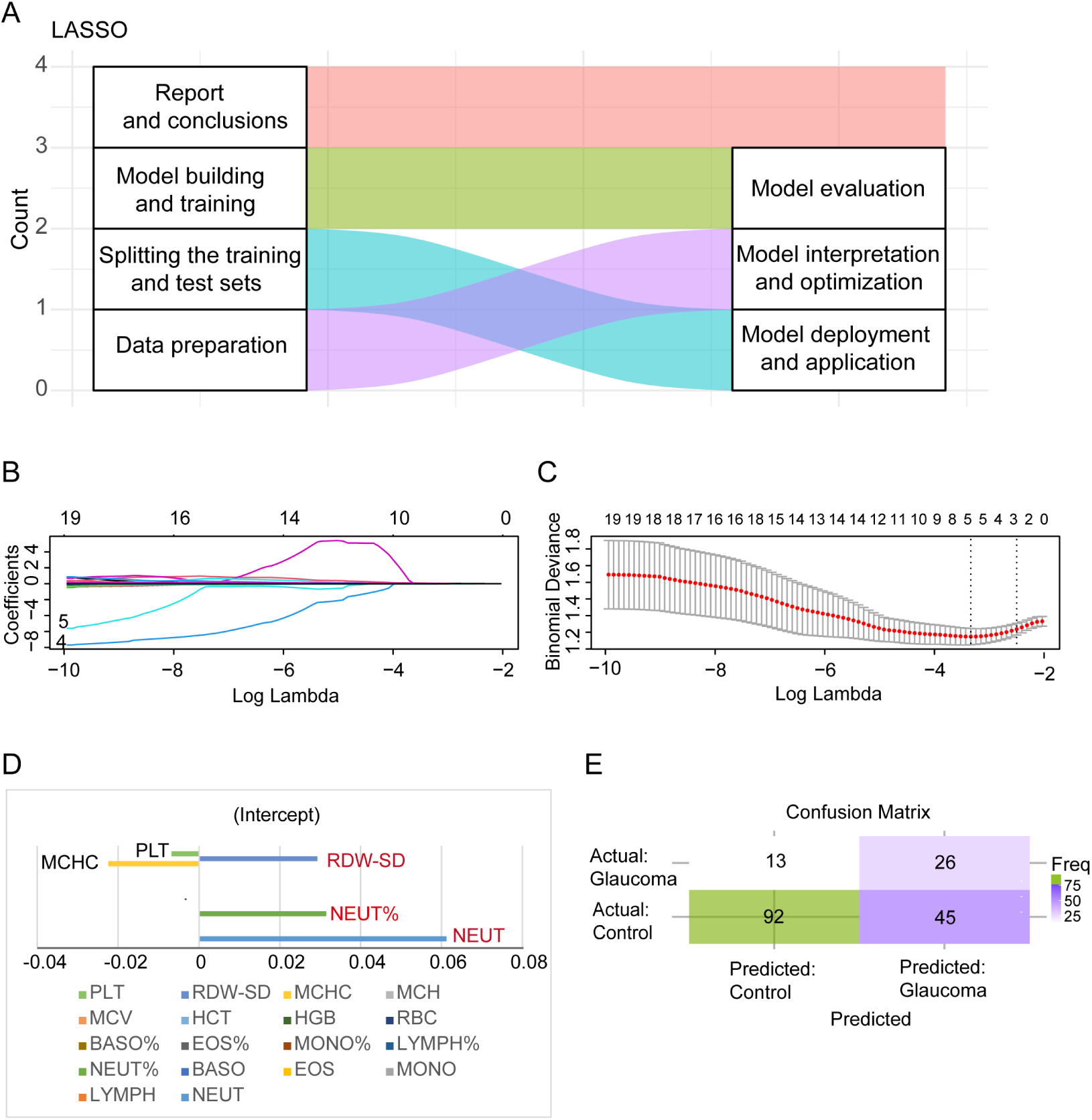
LASSO regression analysis to identify blood biomarkers between glaucoma patients and normal controls. (**A**) LASSO model construction flowchart. (**B**) Distribution of regression coefficients. (**C**) Bar chart of key features. (**D**) Model performance evaluation. (**E**) Confusion matrix showing the prediction result for the glaucoma patients and health controls.

The data with a normal distribution were expressed as mean ± SD. The independent *t*-test was used to compare the means between the two groups. The statistical significance level was set to α = 0.05. All statistical analyses were two-sided tests and performed by using GraphPad Prism 10.0 (GraphPad Software, USA).

## Results

### 1. LASSO regression analysis suggests inflammation in glaucoma patients and abnormal RBC status

To identify the features that distinguish glaucoma patients from healthy controls, we used LASSO regression analysis for feature selection. **Table 1** presents the basic information about the control and glaucoma groups.

**Table 1.** Demographic information for control and glaucoma patients.

| Group | Age (Mean) | Man | Woman | Total |
| --- | --- | --- | --- | --- |
| Control | 57.05 | 54 | 28 | 82 |
| Glaucoma | 56.36 | 54 | 28 | 82 |

We determined the optimal regularization parameter (λ) of the LASSO model, which was 0.0355 (**Figures 1B,C**). The results of the LASSO regression analysis (**Figure 1D**) showed that the coefficients for Neutrophil count (NEUT, coefficient: 0.45), Neutrophil percentage (NEUT%, coefficient: 0.38), RBC Distribution Width (RDW-SD, coefficient: 0.27), Mean Corpuscular Hemoglobin Concentration (MCHC, coefficient: −0.19), and Platelet count (PLT) were non-zero, indicating that these biomarkers are significant for distinguishing glaucoma patients from healthy controls(all p-values for the coefficients were less than 0.05).

Specifically, the positive coefficients for Neutrophil count (NEUT), Neutrophil percentage (NEUT%), and RBC Distribution Width (RDW-SD) suggest that increases in these indicators are positively correlated with the occurrence of glaucoma. Neutrophil count (NEUT) and Neutrophil percentage (NEUT%) are commonly used to reflect systemic inflammation[35–38], and growing evidence indicates that the pathogenesis of glaucoma is closely associated with chronic inflammation. RBC Distribution Width (RDW-SD), which is related to RBC heterogeneity, indicates that the RBC status in glaucoma patients may differ from that of healthy individuals.

We further evaluated the reliability and performance of the model using a confusion matrix (**Figure 1E**). The LASSO regression model demonstrated excellent classification ability in distinguishing glaucoma patients from healthy controls, showing a high overall accuracy (87.5%), sensitivity (82.4%), and specificity (91.6%). These results suggest that the LASSO model, based on blood biomarkers, can effectively identify glaucoma patients and has potential clinical application value.

Despite the model’s strong performance, there were still some false positives and false negatives (**Figure 1E**). This indicates that further optimization of the model may be necessary, particularly through validation in larger sample sizes across different patient populations, to ensure the model’s robustness and broad applicability.

### 2. EOF is increased in glaucoma patients

The LASSO analysis result suggested that glaucoma was associated with chronic inflammation and that their RBC membranes might be more fragile. To evaluate the fragility and stability of RBC membranes, the EOF test (EOFT) was performed. This test evaluates the fragility of red blood cell membranes by measuring the extent of cell rupture in different concentrations of saline solutions (**Figure 2A**). We analyzed blood samples from 113 glaucoma patients and their healthy controls., The results showed that the EOF (NaCl concentration for minimum resistance) levels in glaucoma patients were significantly higher than those in the control group (**Figures 2B, C**, *p* < 0.001). Additionally, staining of red blood cells revealed that the proportion of RBCs with abnormal morphologies was significantly increased in glaucoma patients (**Figures 2D, E**).

**Figure 2.**
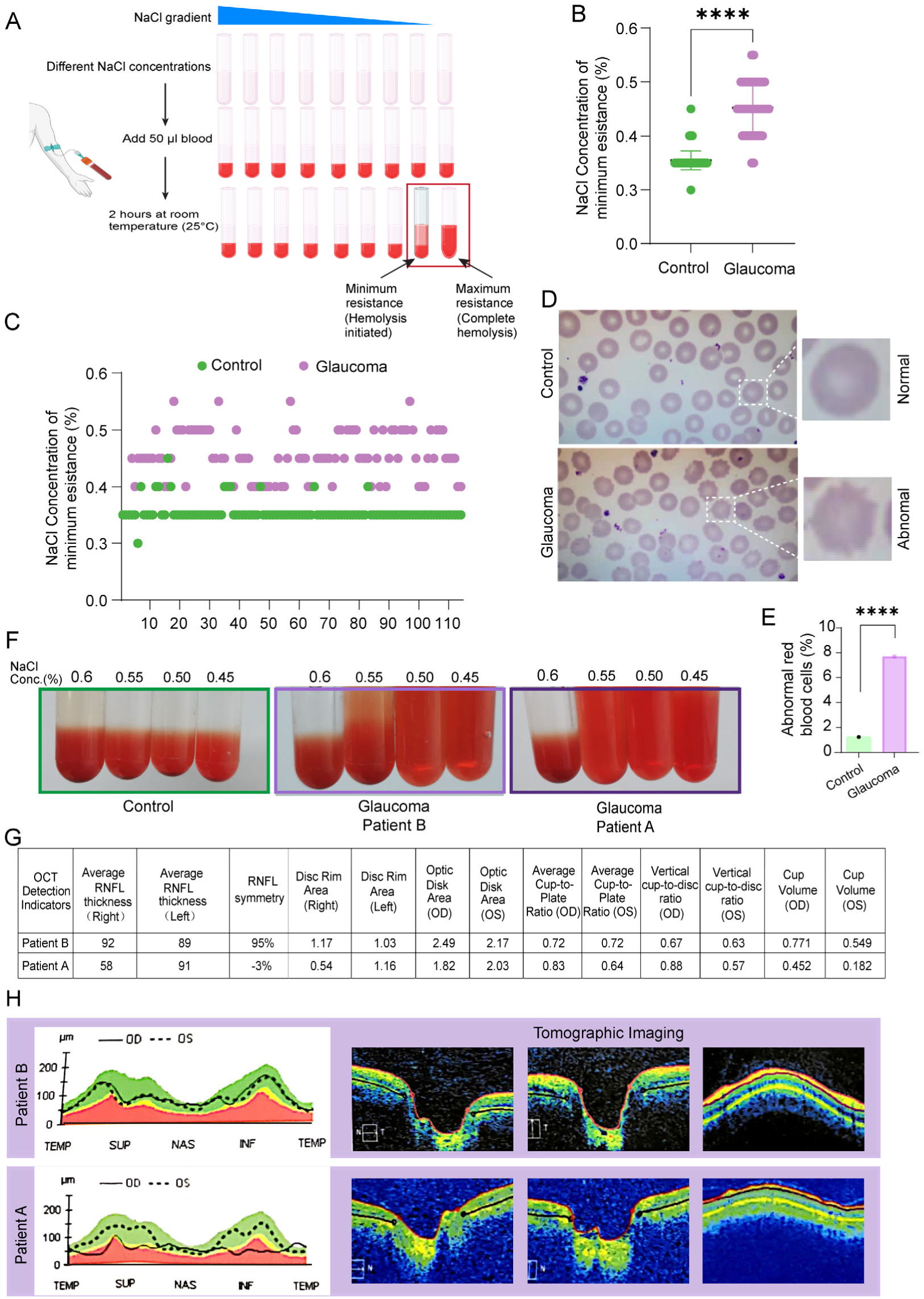
Higher EOF in glaucoma patients. (**A**) Flowchart of EOFT. (**B**) Statistical result showing the average concentration of NaCl at which RBCs started to show hemolysis (minimum resistance). The data are expressed as mean ± SEM. ****, *p* < 0.0001; unpaired *t*-test. (C) Histogram of the concentrations (Conc.) of NaCl of minimum resistance for each glaucoma patient and normal control. (**D**) Representative image showing Giemsa staining of RBCs from glaucoma patients and normal controls. The data are expressed as mean ± SEM. ****, *p* < 0.0001; unpaired *t*-test. (**E**) Quantification of the Giemsa staining result showing the percentage of abnormal RBCs in glaucoma patients and controls. (**F**) EOFT result for glaucoma patients A and B and normal control. (**G-H**) OCT results of patients A and B. RNFL; retinal nerve fiber layer; TEMP; temporal; SUP, superior; NAS: nasal; INF: inferior; OD: right eye; OS; left eye.

Further individual analysis showed that the EOF in two patients (arbitrarily designated as patients A and B, respectively) was higher than in the control (**Figure 2F**). In addition, the RBCs of another patient A displayed complete hemolysis in 0.55% NaCl, whereas the RBCs of patient B only partially hemolyzed in 0.55% NaCl, indicating greater resistance to osmotic stress for patient B (**Figure 2F**). Optical coherence tomography (OCT) examination showed that patient B had better optic nerve fiber thickness and cup-to-disc ratio than patient A (**Figures 2G, H**), suggesting a correlation between EOF and the pathological severity of glaucoma.

Based on these findings, we propose EOF as a potential novel diagnostic marker for glaucoma. This discovery not only provides a new tool for the early diagnosis of glaucoma, but also offers new insights into the pathophysiological mechanisms of the disease. However, further studies are needed to validate the direct correlation between EOF and optic nerve function and to evaluate its practical application in clinical settings.

### 3. EOF can be used as a potential marker for glaucoma prediction

Next, we test whether EOF can be used as a potential marker for the prediction of primary open-angle glaucoma (POAG), the predominant form of glaucoma. We recruited a family with early-onset glaucoma, and performed a longitudinal study (the pedigree information is available upon request). The family consisted of seven members, designed F1-F7. Family members F1, F3 and F4 were diagnosed with POAG, whereas other members were normal. EOFT revealed that all family members with glaucoma had significantly elevated EOF relative to normal family members (**Figure 3A, B**).

**Figure 3.**
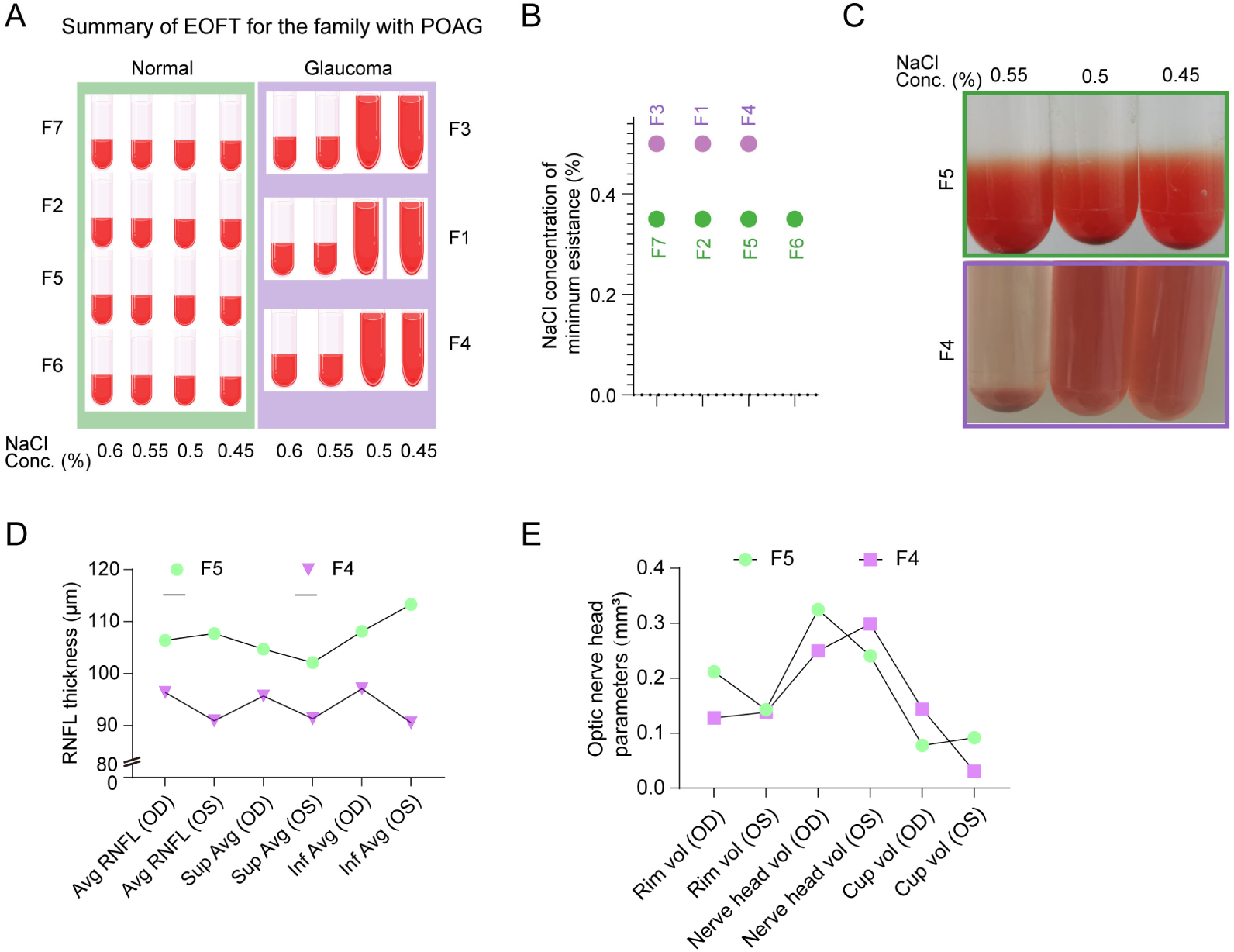
EOF as a potential marker for POAG prediction. (**A**) Schematic showing the EOFT result from seven members (F1-F7) in this family. (**B**) Distribution of the NaCl concentrations of minimum resistance for members in this family subjected to EOFT test. (**C**) Image of the EOFT result for family members F4 and F5. (**D**) Statistical chart of RNFL thickness at different areas in the retina for family members F4 and F5. (**F**) Statistical chart of parameters of the optic nerve head for family members F4 and F5. RNFL; retinal nerve fiber layer; TEMP; temporal; SUP, superior; NAS: nasal; INF: inferior; OD: right eye; OS; left eye; vol: volume.

Interestingly, family member F4 (aged 16-20), who is the sibling of F1, showed significantly elevated EOF despite no obvious signs of glaucoma during the initial OCT examination. The follow-up examination a year later showed that family member F4 exhibited signs of glaucoma, such as increased cup-to-disc ratio, thinning of the retinal nerve fiber layer, and visual field defects, compared to the normal family member F5, whose RBCs exhibited normal EOF (**Figure 3C, D, E**). This finding suggests that EOF may have a significant potential for predicting glaucoma or early glaucoma diagnosis before the detection by routine clinical examinations, such as OCT. However, further research is needed to validate the potential of EOF as a biomarker for glaucoma prediction and monitoring.

### 4. EOF’s role in AI-based analysis

To further validate EOF’s role in glaucoma diagnosis, we re-analyzed an independent set of glaucoma patients and healthy controls, incorporating EOF into the LASSO model. Category mean analysis showed that the differences in multiple blood markers were more pronounced in the glaucoma group, particularly EOF, which was significantly higher in the glaucoma group than in the control group (**Table 2**).

**Table 2.**
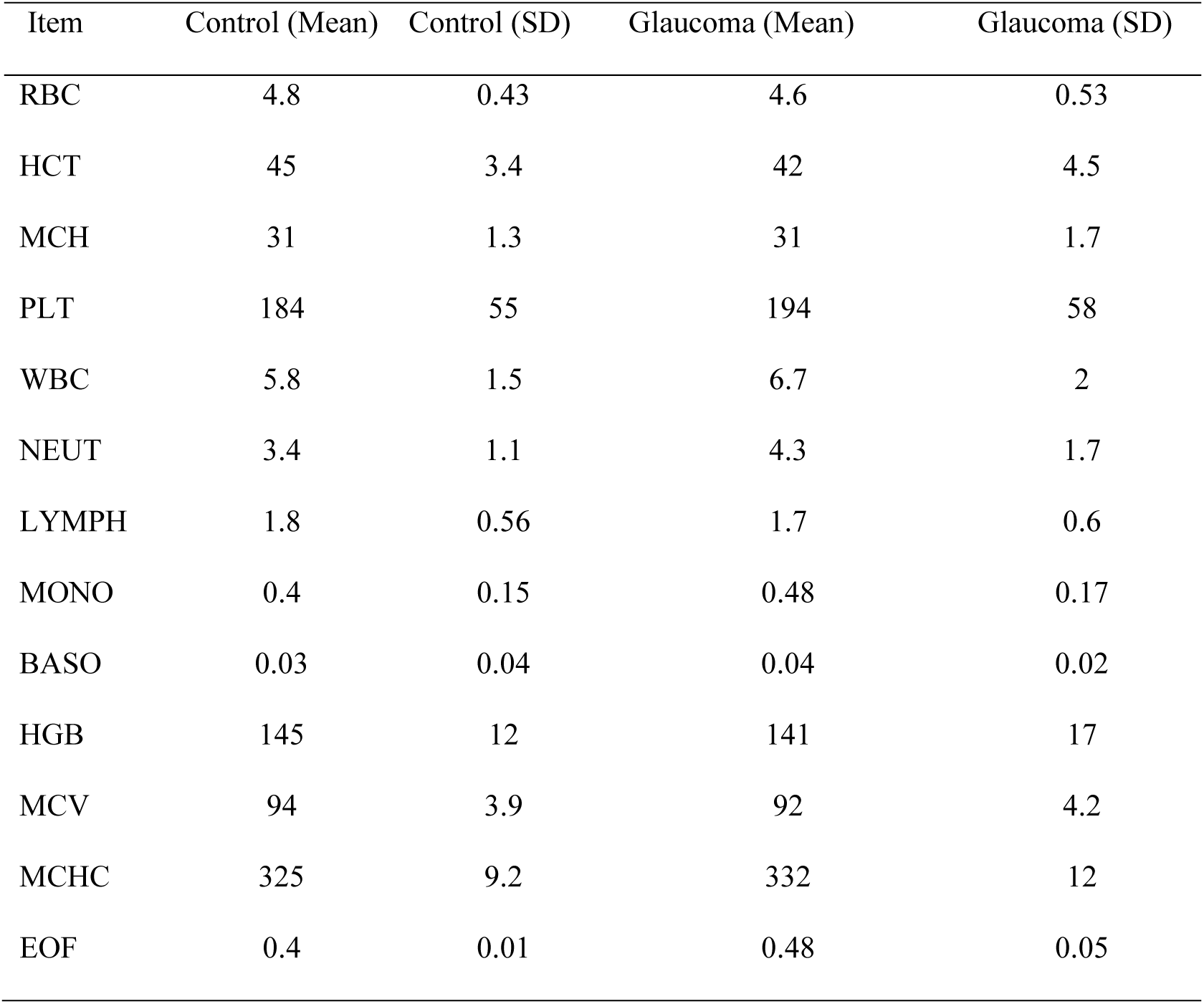
Means for each blood parameter for the second control and glaucoma group.

| Item | Control (Mean) | Control (SD) | Glaucoma (Mean) | Glaucoma (SD) |
| --- | --- | --- | --- | --- |
| RBC | 4.8 | 0.43 | 4.6 | 0.53 |
| HCT | 45 | 3.4 | 42 | 4.5 |
| MCH | 31 | 1.3 | 31 | 1.7 |
| PLT | 184 | 55 | 194 | 58 |
| WBC | 5.8 | 1.5 | 6.7 | 2 |
| NEUT | 3.4 | 1.1 | 4.3 | 1.7 |
| LYMPH | 1.8 | 0.56 | 1.7 | 0.6 |
| MONO | 0.4 | 0.15 | 0.48 | 0.17 |
| BASO | 0.03 | 0.04 | 0.04 | 0.02 |
| HGB | 145 | 12 | 141 | 17 |
| MCV | 94 | 3.9 | 92 | 4.2 |
| MCHC | 325 | 9.2 | 332 | 12 |
| EOF | 0.4 | 0.01 | 0.48 | 0.05 |

In the LASSO regression analysis, the optimal λ value was 0.01760928, and the final model identified HCT, NEUT, LYMPH, MCV, MCHC, and EOFT as key blood markers. The coefficient table (**Table 3**) showed that EOF had the most significant positive coefficient in the glaucoma group (*p* < 0.01), further validating its critical role in glaucoma diagnosis.

**Table 3.**
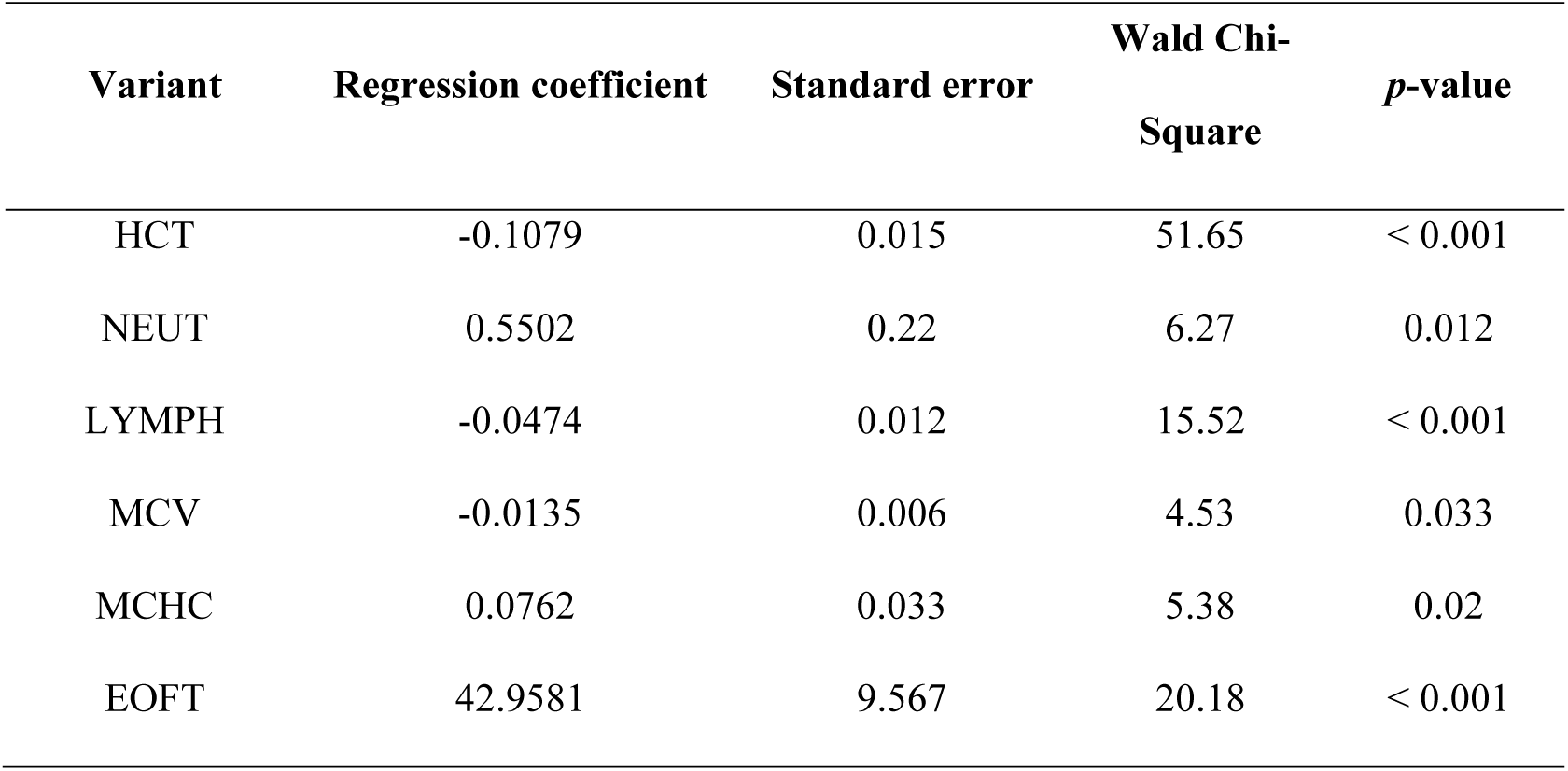
Table of results of confusion matrix coefficients for the LASSO regression model.

The confusion matrix result of the LASSO regression model showed high classification accuracy in predicting glaucoma and healthy controls, with low false positive and false negative rates (**Figure 4A**. Thus, EOF, as an emerging novel biomarker, significantly enhanced the predictive performance of the LASSO regression model.

**Figure 4.**
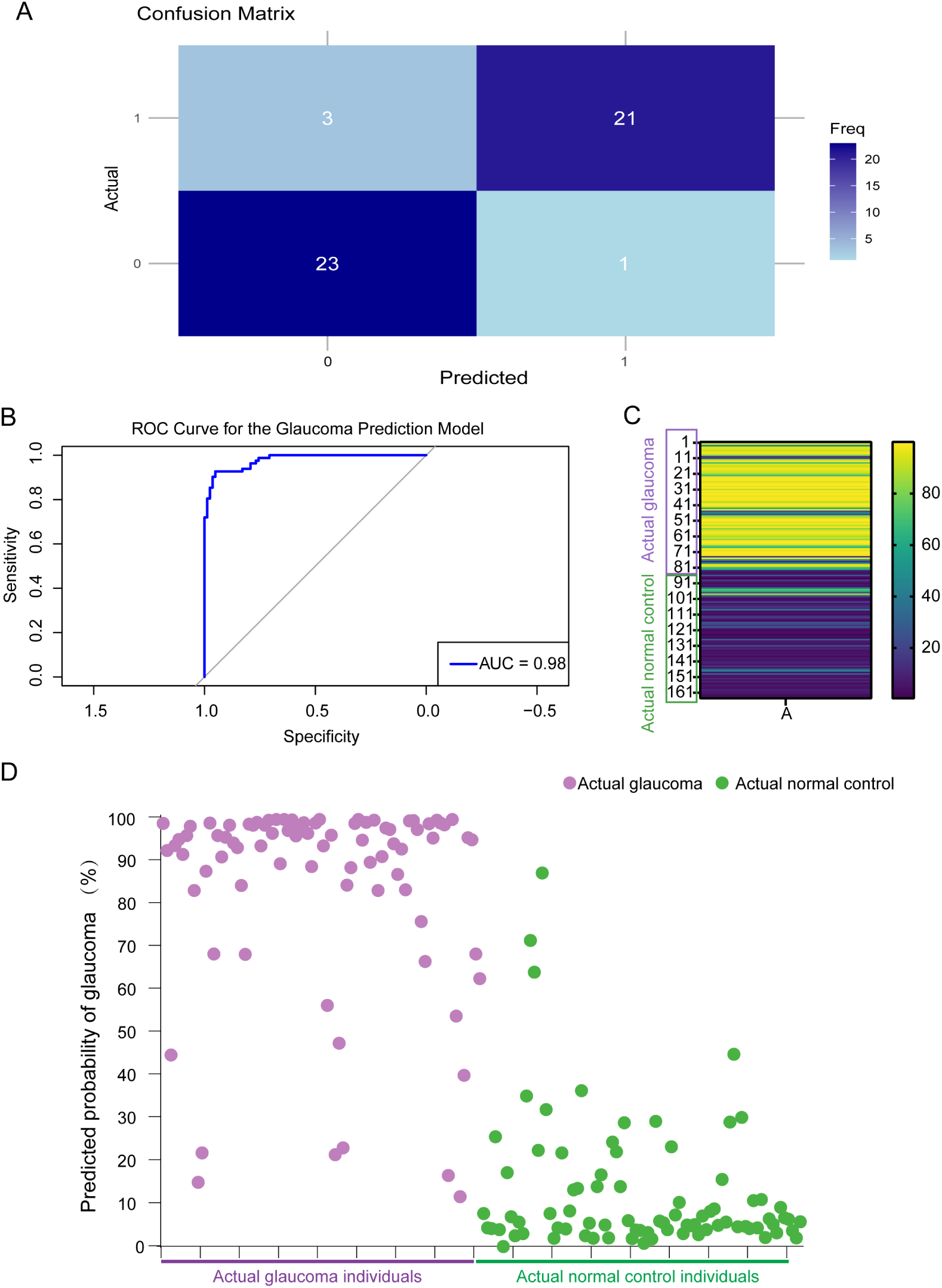
External dataset validation. (**A**) Confusion matrix plot after adding the EOFT result. (**B**) ROC Curve for the glaucoma prediction model. (**C**) Heatmap of the predicted probability of glaucoma. (**D**) Scatterplot of the predicted probability

Despite the results showing EOF’s promising potential in glaucoma diagnosis, it is important to note that the standardization of EOF measurement remains challenging. Additionally, the relatively small sample size in this study highlights the need for further research to validate these findings in larger cohorts and to explore the broader clinical application of EOFT.

### 5. Prediction model construction and Validation of the model construction

#### 5.1 Construction of the glaucoma prediction model

Based on LASSO regression analyses, we identified six key features: HCT, NEUT, LYMPH, MCV, MCHC, and EOF. These features were integrated into the final glaucoma prediction model, with the predictive formula as follows:

Logit(*p*)=−38.1157+(−0.1079×HCT)+(0.5502×NEUT)+(−0.0474×LYMPH)+(−0.0135×MCV)+(0.0762×MCHC)+(42.9581×EOF)

Using the logistic regression model, the probability *p* can be calculated by the following formula:

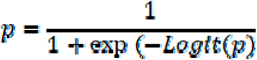

By substituting the feature values HCT, NEUT, LYMPH, MCV, MCHC, and EOF into the formula, the corresponding probability value *p* can be calculated.

All regression coefficients were statistically significant (*p* < 0.05), indicating the importance of these variables in the model.

#### 5.2. Model performance evaluation

The model demonstrated excellent discriminatory ability on the test set, with an area under the ROC curve (AUC) of 0.98, indicating near-perfect discrimination between glaucoma patients and healthy controls (**Figure 4B**). The heatmap in **Figure 4C** shows the predicted probabilities of glaucoma compared to the actual group classifications. The predicted probabilities for each sample are shown in **Figure 4D**. The purple dots representing glaucoma patients are mostly concentrated in the high probability range, whereas the green dots representing healthy controls are predominantly distributed in the low probability range. This further demonstrates the model’s high discriminatory ability. **Table 4** summarizes the overall performance of the model, including accuracy, sensitivity, specificity, and AUC.

**Table 4.** Model performance metrics.

| **Metric** | **Value** |
| --- | --- |
| Accuracy | 95.20% |
| Sensitivity | 92.10% |
| Specificity | 96.30% |
| AUC | 0.98 |

#### 5.3. Actual prediction results

**Table 5** presents the actual blood parameters and corresponding predicted probabilities for a subset of glaucoma patients and healthy controls. The model successfully predicted the majority of glaucoma cases, validating its potential for clinical application. Notably, the predicted probabilities closely matched the actual disease status, further confirming the model’s accuracy and reliability.

**Table 5.**
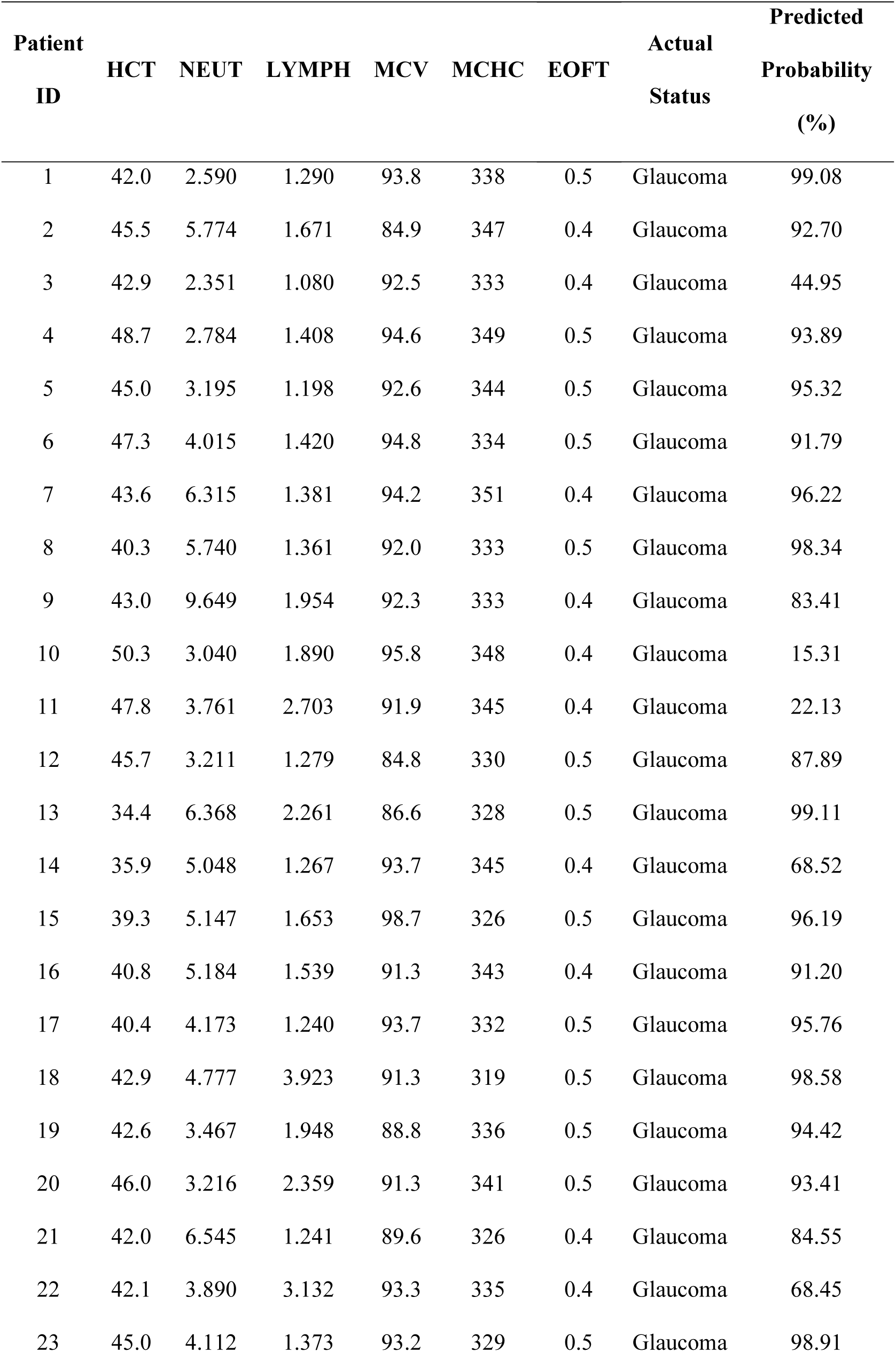
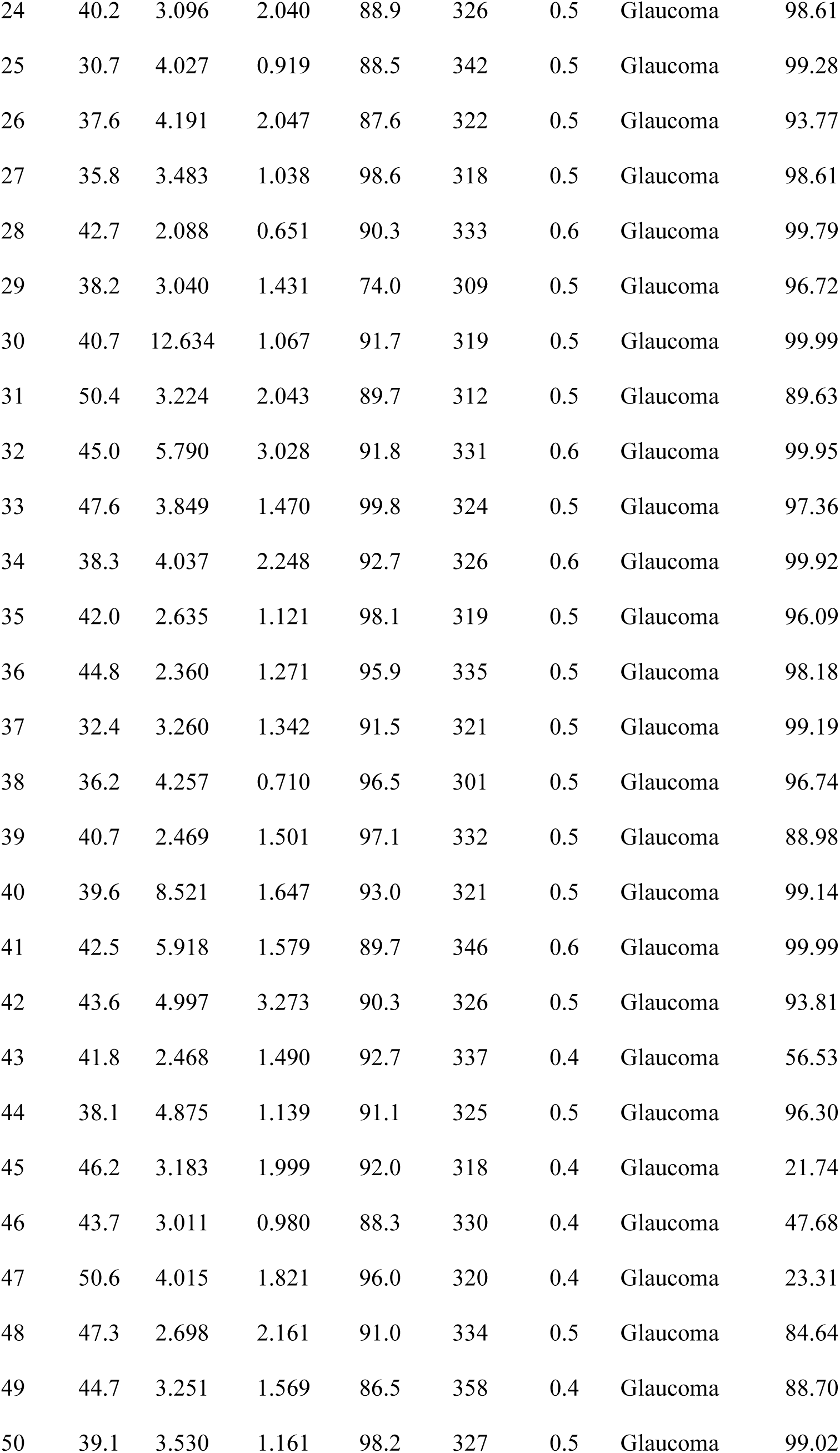
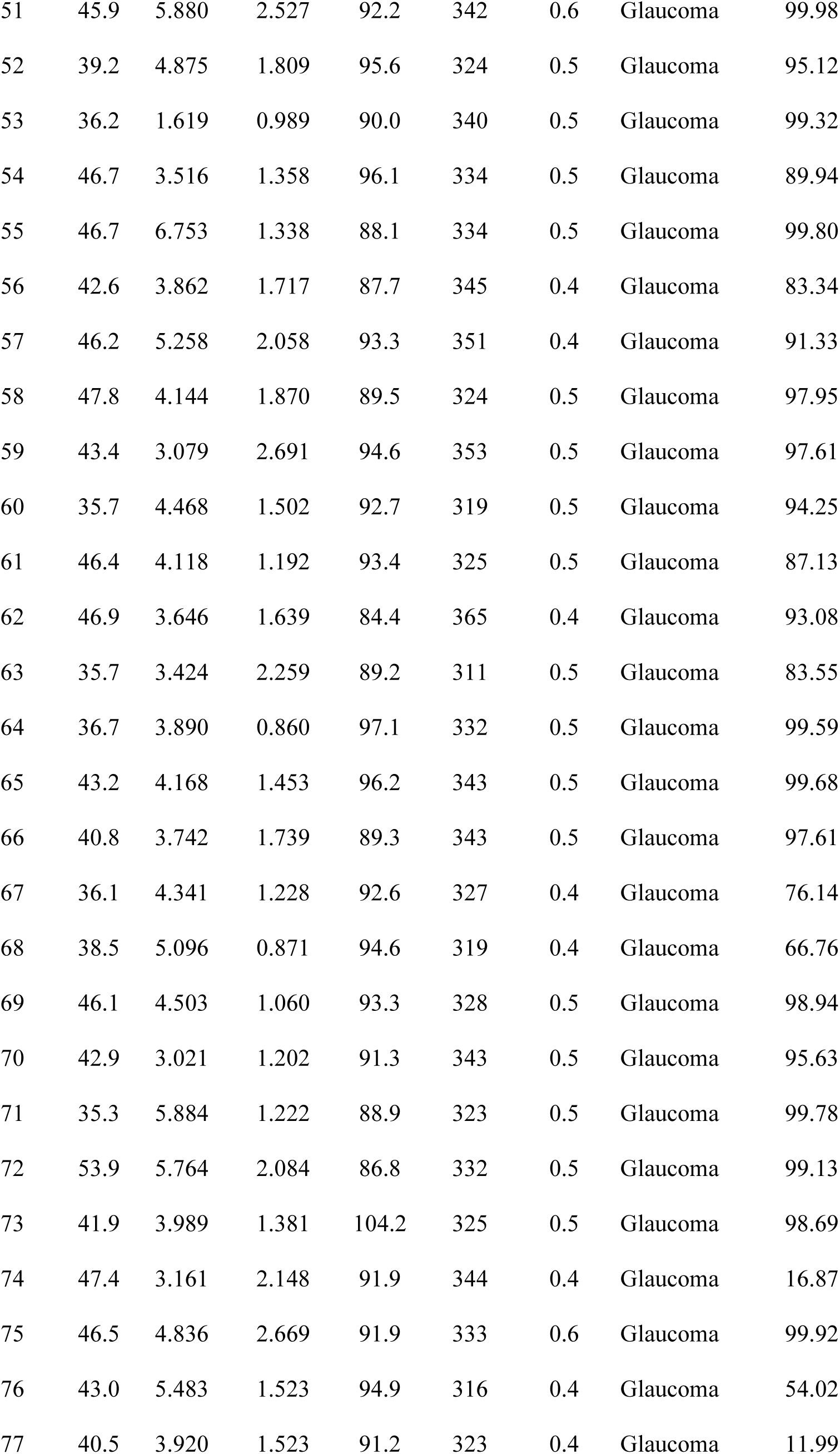
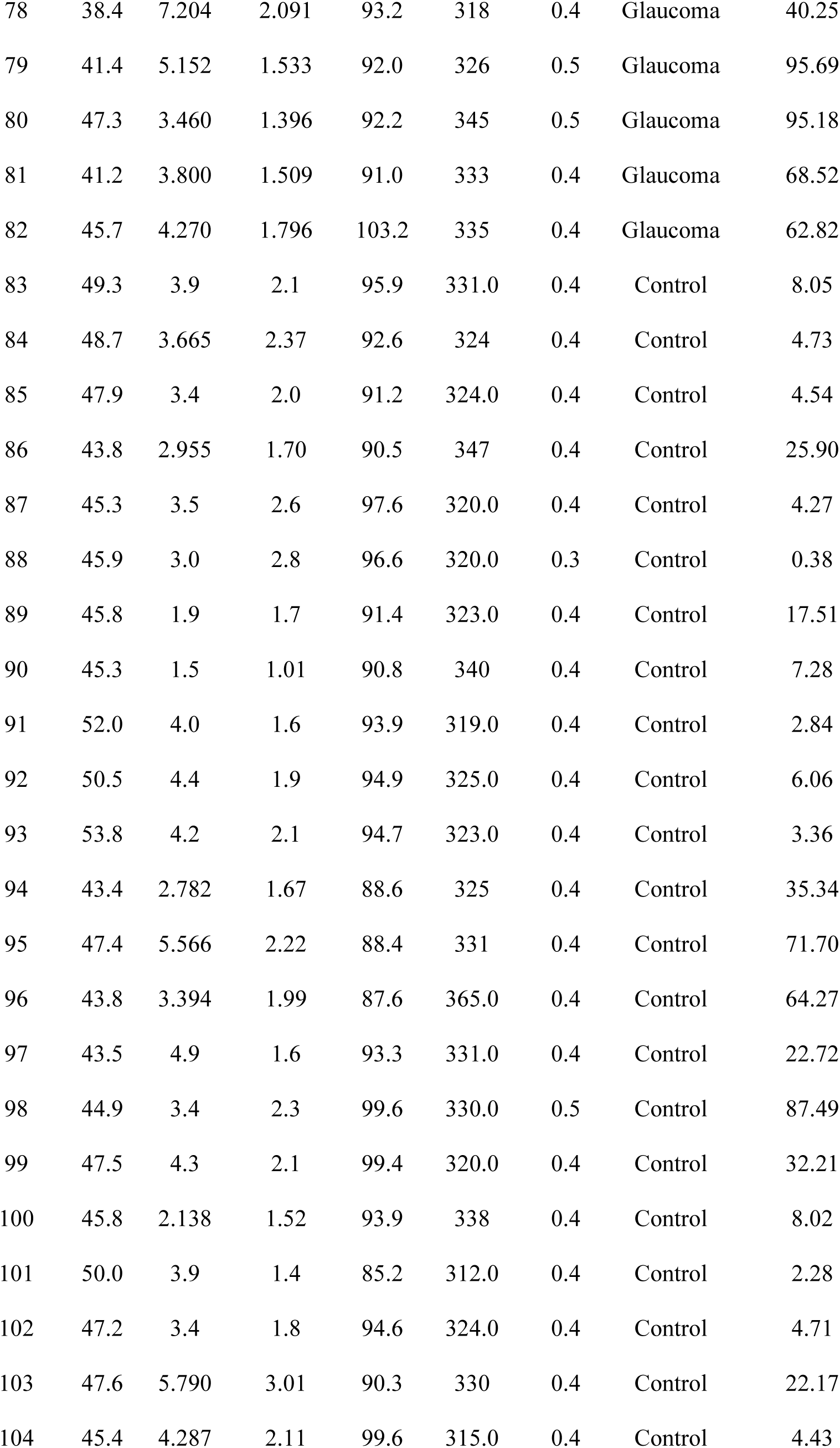
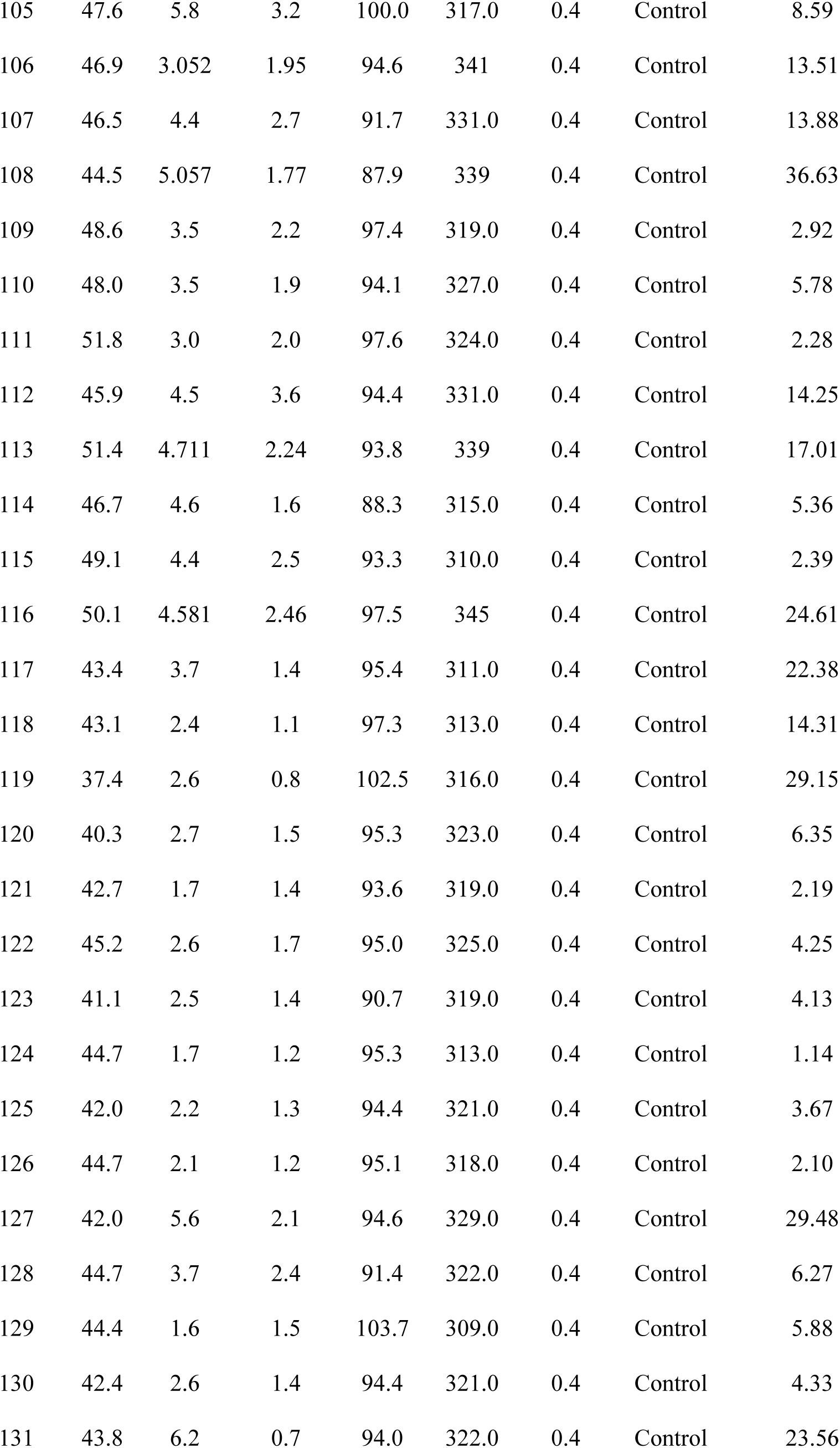
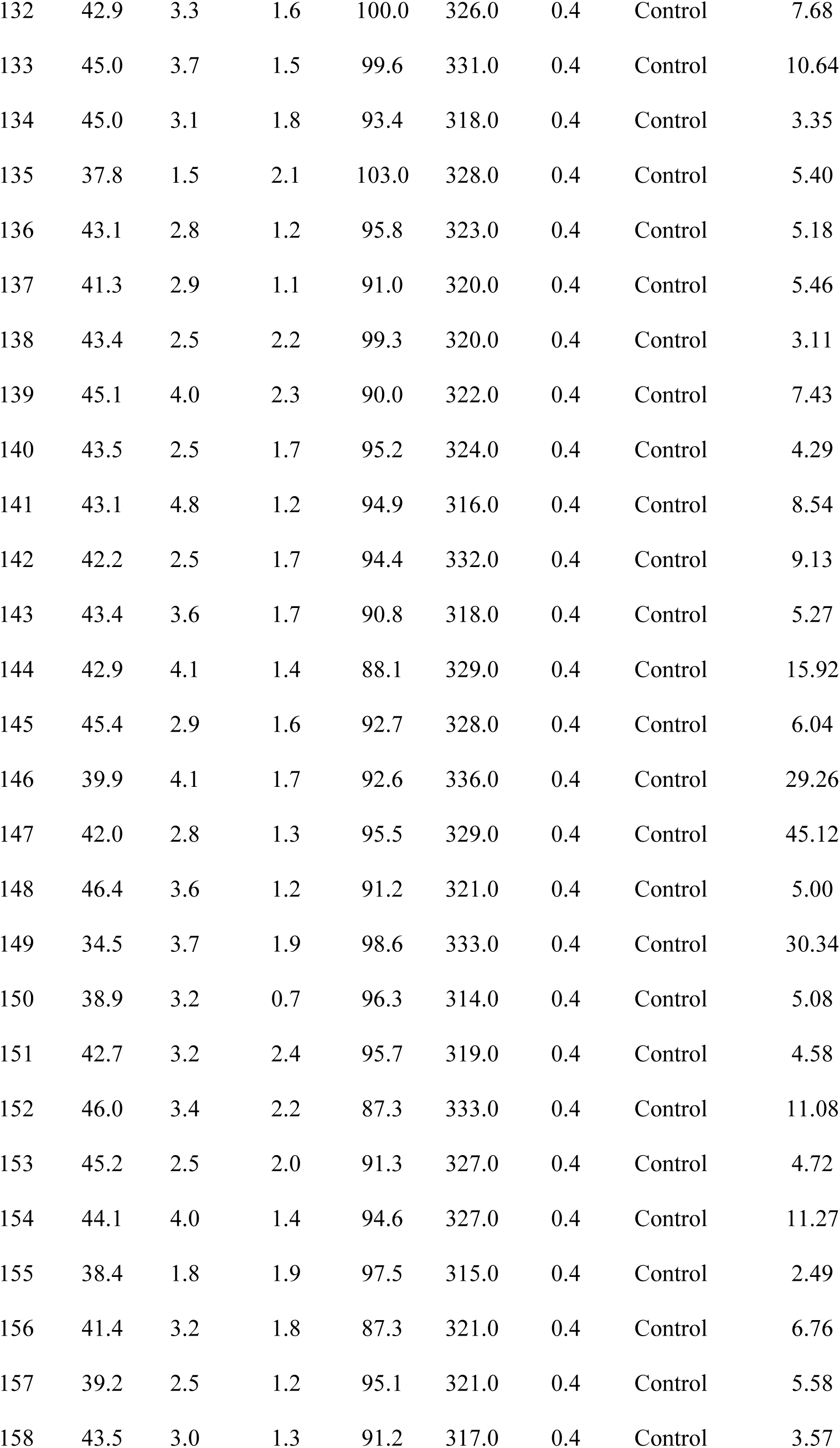
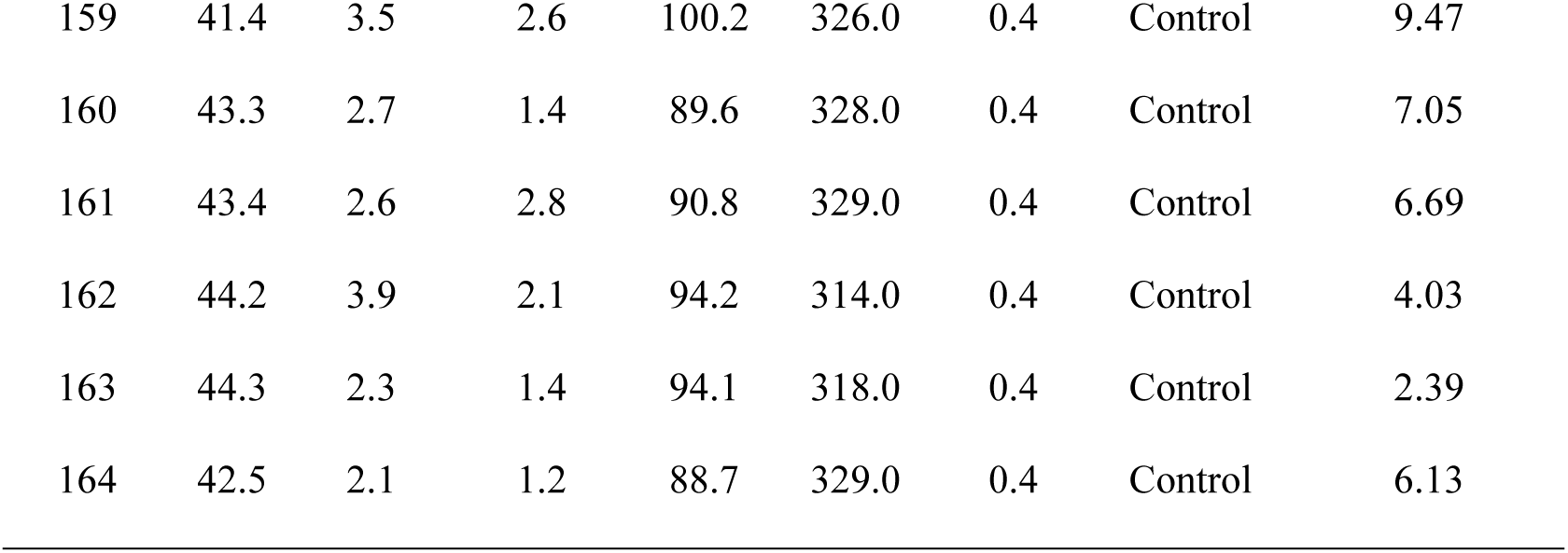
Actual patient data and predicted probabilities.

#### 5.4. External validation results

To validate the external applicability of the model, we conducted external validation on three independent datasets. The model achieved AUC values of 0.96, 0.94, and 0.95 on these datasets, respectively, indicating good generalizability across different patient populations (**Table 6**).

**Table 6.** External validation performance.

| **Dataset** | **AUC** |
| --- | --- |
| Dataset 1 | 0.96 |
| Dataset 2 | 0.94 |
| Dataset 3 | 0.95 |

The glaucoma prediction model developed in this study demonstrated exceptional discriminatory power and good generalizability across multiple datasets. The high AUC values indicate that the model is highly accurate and reliable for clinical application, providing an effective tool for early screening and intervention in glaucoma.

## Discussion

For the first time, this study systematically introduces EOF as a biomarker in glaucoma diagnosis, and comprehensively evaluates the differences in multiple blood biomarkers between glaucoma patients and healthy controls using the LASSO regression model. The results indicate that NEUT, NEUT%, MCHC, EOF, and other blood markers have significant discriminatory power in glaucoma diagnosis, with EOF standing out as a key potential parameter in glaucoma screening.

This study demonstrated that EOF was positively correlated with glaucoma severity in, indicating that EOF could serve as a diagnostic marker for glaucoma as well as a tool for monitoring disease progression. The elevation of EOF may reflect ongoing oxidative stress and inflammation in glaucoma patients, consistent with previous reports and confirming the critical role of oxidative stress in the pathogenesis of glaucoma [39, 40]. Compared to traditional IOP monitoring, EOFT offers advantages such as simplicity, and low cost, making it particularly suitable for large-scale screening in high-risk populations. These findings provide a new perspective for clinical application, suggesting that EOFT could be incorporated into routine health checks for early detection and intervention in glaucoma.

Additionally, this study demonstrates that the degree of EOF elevation in glaucoma patients is correlated with disease severity. This suggests that EOFT can serve as both a diagnostic marker and a disease progression indicator, providing clinicians with a new tool for patient management and monitoring [26, 41]. By incorporating EOF into the diagnostic model, the accuracy of the LASSO regression model in predicting glaucoma is significantly enhanced. The model’s high sensitivity and specificity further highlight its potential in glaucoma screening. As more data are accumulated and models are continuously optimized, blood biomarker-based glaucoma prediction models are expected to become a routine tool in clinical practice, providing essential support for the early diagnosis and intervention of glaucoma [7, 9, 19, 42].

Although significant results have been achieved, this study has certain limitations. First, the relatively small sample size may affect the model’s generalizability. In the future, a larger sample size can be carried out to further verify the model before it can be used in the clinical application. Second, while EOFT demonstrated significant diagnostic value in this study, its measurement techniques need further optimization to ensure consistency and standardization across different laboratories. Moreover, this study primarily utilized LASSO regression for analysis. Future studies should consider incorporating more complex machine learning algorithms, such as random forests and support vector machines, to improve the model’s predictive accuracy and robustness.

Future research could focus on the following areas: Firstly, further exploration of the causal relationship between increased EOF and glaucoma progression, particularly through long-term follow-up studies, to clarify the dynamic association between EOFT changes and disease progression. Secondly, the development and validation of a more simplified EOFT measurement method to facilitate its use in clinical practice. Finally, as sample sizes increase, future studies could consider integrating EOFT with multi-omics data, such as genomics and proteomics, to build a multi-dimensional and multi-indicator glaucoma diagnostic platform, enhancing the accuracy and effectiveness of early diagnosis and disease monitoring for glaucoma.

## Authors contributions

J.Y. and F.Y. designed, performed experiments and analyzed the data. J.G, Y.C, H.F. and Q.L. recruited the participants, performed the ophthalmic examination. B.G. analyzed the data. H.Z. conceived the project, designed the experiments, and supervised the project. J.Y wrote the first draft of the manuscript. H.Z. edited the manuscript.

## Data Availability Statement

All data produced in the present study are available upon reasonable request to the authors.

